## Supplemental Tables S1 - S9 for "Does behavior mediate the effect of weather on SARS-CoV-2 transmission? Evidence from cell-phone data"

**COVID-Weather analysis supplement**

**Supplemental Table S1. Detailed regression results between control variables and categorical weather variables on time indoors away-from-home, and 12-day lagged hospitalizations**

|  | **Daily county-mean percent of time spent indoors, away from home** | | | **12-day lagged hospitalization admissions (mean-centered by county-season)** | | |
| --- | --- | --- | --- | --- | --- | --- |
| **Control variables** ^a^ | **β** | **95 % CI** | **p-value** | **β** | **95 % CI** | **p-value** |
| **Weekend/holiday (yes/no)** |  |  |  |  |  |  |
| All Seasons ^b^ | -0.98 | -1.04 - -0.91 | <0.001* | -0.22 | -0.56 – 0.12 | 0.201 |
| Spring | -1.22 | -1.40 - -1.04 | <0.001* | 0.06 | -0.51 – 0.63 | 0.835 |
| Summer | -0.97 | -1.04 - -0.91 | <0.001* | 0.28 | -0.03 – 0.60 | 0.080 |
| Fall | -1.18 | -1.28 - -1.09 | <0.001* | -0.24 | -1.09 – 0.62 | 0.584 |
| Winter | -1.04 | -1.17 - -0.91 | <0.001* | -0.67 | -1.49 – 0.15 | 0.109 |
| **Stay-at-home order (yes/no)** |  |  |  |  |  |  |
| All Seasons ^b^ | -0.39 | -0.53 - -0.26 | <0.001* | 0.19 | -0.32 – 0.69 | 0.469 |
| Spring | -0.31 | -0.55 - -0.08 | 0.009* | 0.28 | -0.26 – 0.82 | 0.305 |
| Summer | NA | NA | NA | NA | NA | NA |
| Fall | NA | NA | NA | NA | NA | NA |
| Winter | NA | NA | NA | NA | NA | NA |
| **Rising Colorado hospitalizations (yes/no)** |  |  |  |  |  |  |
| All Seasons ^b^ | -0.11 | -0.19 – -0.03 | 0.005* | 1.15 | 0.81 – 1.49 | <0.001* |
| Spring | -0.30 | -0.49 - -0.10 | 0.004* | 1.17 | 0.58 – 1.76 | <0.001* |
| Summer | 0.07 | -0.05 – 0.18 | 0.253 | 0.27 | -0.08 – 0.62 | 0.131 |
| Fall | -0.01 | -0.14 – 0.13 | 0.904 | 2.50 | 1.55 – 3.45 | <0.001* |
| Winter | -0.31 | -0.50 - -0.12 | 0.001* | 1.58 | 0.61 – 2.55 | 0.001* |
| **Categorical weather conditions** ^c^ | **β** | **95 % CI** | **p-value** | **β** | **95 % CI** | **p-value** |
| **Minimum temperature** |  |  |  |  |  |  |
| All season ^b^ |  |  |  |  |  |  |
| <-0.5 vs. mid | 0.09 | 0.01 – 0.17 | 0.028* | 0.14 | -0.25 – 0.53 | 0.490 |
| >0.5 vs. mid | -0.09 | -0.16 - -0.01 | 0.020* ^d^ | -0.41 | -0.78 - -0.03 | 0.034* ^d^ |
| Spring |  |  |  |  |  |  |
| <-0.5 vs. mid | 0.30 | 0.11 – 0.48 | 0.002* | -0.18 | -0.78 – 0.42 | 0.557 |
| >0.5 vs. mid | -0.19 | -0.41 – 0.04 | 0.099 | -0.23 | -0.96 – 0.49 | 0.526 |
| Summer |  |  |  |  |  |  |
| <-0.5 vs. mid | -0.08 | -0.16 - -0.00 | 0.039* | -0.35 | -0.77 – 0.07 | 0.102 |
| >0.5 vs. mid | -0.09 | -0.16 - -0.03 | 0.004* | -0.14 | -0.47 – 0.19 | 0.394 |
| Fall |  |  |  |  |  |  |
| <-1 vs. mid | -0.05 | -0.18 – 0.08 | 0.452 | 0.61 | -0.56 – 1.78 | 0.306 |
| >1 vs. mid | -0.08 | -0.20 – 0.04 | 0.210 | -0.03 | -1.12 – 1.06 | 0.959 |
| Winter |  |  |  |  |  |  |
| <-1 vs. mid | -0.32 | -0.47 – -0.17 | <0.001* | 0.26 | -0.69 – 1.21 | 0.589 |
| >1 vs. mid | 0.04 | -0.13 – 0.22 | 0.640 | 1.19 | -0.01 – 2.40 | 0.053 |
| **Maximum temperature** |  |  |  |  |  |  |
| All season ^b^ |  |  |  |  |  |  |
| <-1 vs. mid | 0.08 | -0.00 – 0.17 | 0.060 | -0.19 | -0.62 – 0.23 | 0.374 |
| >1 vs. mid | -0.11 | -0.20 – 0.01 | 0.024* | -0.28 | -0.74 – 0.19 | 0.241 |
| Spring |  |  |  |  |  |  |
| <-1 vs mid | 0.23 | 0.03 – 0.44 | 0.028* ^d^ | -0.73 | -1.41 - -0.04 | 0.037* ^d^ |
| >1 vs mid | -0.12 | -0.42 – 0.18 | 0.448 | -0.62 | -1.58 – 0.33 | 0.201 |
| Summer |  |  |  |  |  |  |
| <-1 vs. mid | -0.05 | -0.14 – 0.04 | 0.303 | -0.48 | -0.94 - -0.02 | 0.039* |
| >1 vs. mid | -0.08 | -0.16 – -0.00 | 0.954 | -0.18 | -0.59 – 0.23 | 0.390 |
| Fall |  |  |  |  |  |  |
| <-1 vs. mid | -0.09 | -0.21 – 0.03 | 0.145 | 0.32 | -0.78 – 1.41 | 0.567 |
| >1 vs. mid | -0.13 | -0.26 - -0.01 | 0.041* ^d^ | -0.28 | -1.42 – 0.86 | 0.632 |
| Winter |  |  |  |  |  |  |
| <-1 vs. mid | -0.23 | -0.37 - -0.08 | 0.003* | 0.40 | -0.59 – 1.39 | 0.431 |
| >1 vs. mid | 0.13 | -0.03 – 0.30 | 0.120 | 0.23 | -0.91 – 1.37 | 0.691 |
| **Minimum Relative Humidity** |  |  |  |  |  |  |
| All season ^b^ |  |  |  |  |  |  |
| <-1 vs. mid | -0.09 | -0.21 – 0.02 | 0.100 | -0.17 | -0.73 – 0.39 | 0.557 |
| >1 vs. mid | -0.06 | -0.15 – 0.03 | 0.209 | -0.27 | -0.72 – 0.18 | 0.238 |
| Spring |  |  |  |  |  |  |
| <-0.5 vs. mid | -0.35 | -0.52 - -0.18 | <0.001* | -0.03 | -0.60 – 0.55 | 0.926 |
| >1 vs. mid | -0.22 | -0.44 - -0.00 | 0.048* | -0.65 | -1.40 – 0.10 | 0.089 |
| Summer |  |  |  |  |  |  |
| <-0.5 vs. mid | 0.02 | -0.04 – 0.08 | 0.435 | 0.12 | -0.19 – 0.44 | 0.439 |
| >1 vs. mid | -0.01 | -0.09 – 0.08 | 0.880 | 0.02 | -0.43 – 0.47 | 0.930 |
| Fall |  |  |  |  |  |  |
| <-0.5 vs. mid | -0.03 | -0.13 – 0.06 | 0.474 | -0.05 | -0.90 – 0.79 | 0.899 |
| >1 vs. mid | -0.19 | -0.33 - -0.05 | 0.008* | 0.39 | -0.86 – 1.64 | 0.540 |
| Winter |  |  |  |  |  |  |
| <-1 vs. mid | 0.23 | 0.04 – 0.43 | 0.018* | -1.04 | -2.36 – 0.27 | 0.121 |
| >1 vs. mid | -0.27 | -0.43 - -0.11 | 0.001* | -0.01 | -1.11 – 1.09 | 0.985 |
| **Maximum relative humidity** |  |  |  |  |  |  |
| All Season ^b^ |  |  |  |  |  |  |
| <-1 vs. mid | -0.11 | -0.19 - -0.02 | 0.015* | -0.42 | -0.85 – 0.12 | 0.057 |
| >1 vs. mid | -0.03 | -0.11 – 0.05 | 0.458 | -0.36 | -0.76 – 0.04 | 0.081 |
| Spring |  |  |  |  |  |  |
| <-1 vs. mid | -0.57 | -0.78 - -0.37 | <0.001* | -0.14 | -0.84 – 0.57 | 0.706 |
| >1 vs. mid | -0.14 | -0.33 – 0.05 | 0.153 | -0.14 | -0.80 – 0.53 | 0.683 |
| Summer |  |  |  |  |  |  |
| <-0.5 vs. mid | 0.06 | -0.00 – 0.12 | 0.072 | 0.01 | -0.31 – 0.33 | 0.963 |
| >1 vs. mid | 0.04 | -0.04 – 0.12 | 0.348 | 0.00 | -0.42 – 0.42 | 0.999 |
| Fall |  |  |  |  |  |  |
| <-0.5 vs. mid | 0.02 | -0.08 – 0.12 | 0.661 | -0.08 | -0.96 – 0.80 | 0.856 |
| >1 vs. mid | -0.03 | -0.15 – 0.08 | 0.570 | -0.02 | -1.07 – 1.04 | 0.974 |
| Winter |  |  |  |  |  |  |
| <-1 vs. mid | 0.23 | 0.05 – 0.41 | 0.014* | -1.00 | -2.23 – 0.24 | 0.113 |
| >1 vs. mid | -0.25 | -0.40 - -0.11 | 0.001* | -0.45 | -1.44 – 0.54 | 0.376 |
| **Minimum absolute humidity** |  |  |  |  |  |  |
| All Season ^b^ |  |  |  |  |  |  |
| <-1 vs. mid | 0.14 | 0.05 – 0.24 | 0.004* | 0.08 | -0.40 – 0.56 | 0.740 |
| >1 vs. mid | -0.09 | -0.18 – 0.00 | 0.053* | -0.31 | -0.77 – 0.15 | 0.183 |
| Spring |  |  |  |  |  |  |
| <-1 vs mid | -0.03 | -0.26 – 0.20 | 0.813 | 0.45 | -0.33 – 1.22 | 0.259 |
| >1 vs mid | -0.38 | -0.63 - -0.13 | 0.003* | 0.07 | -0.76 – 0.90 | 0.865 |
| Summer |  |  |  |  |  |  |
| <-1 vs. mid | 0.07 | -0.02 – 0.15 | 0.133 | -0.03 | -0.48 – 0.41 | 0.892 |
| >1 vs. mid | -0.10 | -0.17 - -0.02 | 0.011* | -0.16 | -0.55 – 0.22 | 0.401 |
| Fall |  |  |  |  |  |  |
| <-1 vs. mid | 0.18 | 0.04 – 0.33 | 0.014* | 0.68 | -0.66 – 2.01 | 0.320 |
| >1 vs. mid | 0.07 | -0.07 – 0.20 | 0.349 | 0.07 | -1.14 – 1.29 | 0.910 |
| Winter |  |  |  |  |  |  |
| <-1 vs. mid | 0.22 | 0.03 – 0.40 | 0.017* ^d^ | -1.28 | -2.50 - -0.07 | 0.039* ^d^ |
| >1 vs. mid | -0.12 | -0.32 – 0.07 | 0.209 | -0.43 | -1.70 – 0.84 | 0.509 |
| **Maximum absolute humidity** |  |  |  |  |  |  |
| All Season ^b^ |  |  |  |  |  |  |
| <-1 vs. mid | -0.72 | -0.87 – -0.57 | <0.001* | 0.07 | -0.37 – 0.50 | 0.763 |
| >1 vs. mid | -0.11 | -0.18 – -0.04 | 0.002* | -0.11 | -0.58 – 0.36 | 0.657 |
| Spring |  |  |  |  |  |  |
| <-1 vs mid | 0.05 | -0.17 – 0.28 | 0.649 | -1.00 | -1.74 – -0.26 | 0.008* |
| >1 vs mid | -0.15 | -0.46 – 0.15 | 0.312 | -0.12 | -1.07 – 0.84 | 0.810 |
| Summer |  |  |  |  |  |  |
| <-1 vs. mid | 0.05 | -0.03 – 0.12 | 0.201 | 0.01 | -0.38 – 0.40 | 0.963 |
| >1 vs. mid | -0.03 | -0.11 – 0.04 | 0.344 | 0.00 | -0.38 – 0.38 | 0.993 |
| Fall |  |  |  |  |  |  |
| <-1 vs. mid | -0.09 | -0.23 – 0.06 | 0.251 | 1.27 | -0.04 – 2.57 | 0.058 |
| >1 vs. mid | -0.03 | -0.16 – 0.09 | 0.611 | 0.23 | -0.92 – 1.37 | 0.698 |
| Winter |  |  |  |  |  |  |
| <-1 vs. mid | -0.13 | -0.28 – 0.03 | 0.106 | 0.05 | -0.99 – 1.08 | 0.931 |
| >1 vs. mid | -0.06 | -0.31 – 0.18 | 0.619 | 0.05 | -1.56 – 1.67 | 0.950 |
| **Wind Speed** |  |  |  |  |  |  |
| All Season ^b^ |  |  |  |  |  |  |
| <-1 vs. mid | -0.01 | -0.11 – 0.08 | 0.806 | -0.09 | -0.57 – 0.40 | 0.730 |
| >1 vs. mid | 0.03 | -0.06 – 0.12 | 0.469 | 0.39 | -0.06 – 0.84 | 0.093 |
| Spring |  |  |  |  |  |  |
| <-1 vs mid | -0.04 | -0.27 – 0.19 | 0.753 | 0.13 | -0.64 – 0.90 | 0.745 |
| >1 vs mid | 0.10 | -0.12 – 0.32 | 0.360 | 0.03 | -0.70 – 0.76 | 0.930 |
| Summer |  |  |  |  |  |  |
| <-1 vs. mid | -0.00 | -0.09 – 0.09 | 0.950 | -0.09 | -0.57 – 0.40 | 0.725 |
| >1 vs. mid | -0.08 | -0.16 – 0.01 | 0.075 | -0.40 | -0.83 – 0.04 | 0.074 |
| Fall |  |  |  |  |  |  |
| <-1 vs. mid | -0.08 | -0.22 – 0.06 | 0.254 | -0.03 | -1.26 – 1.21 | 0.966 |
| >1 vs. mid | -0.16 | -0.29 - -0.03 | 0.018* | 1.11 | -0.08 – 2.29 | 0.067 |
| Winter |  |  |  |  |  |  |
| <-1 vs. mid | 0.15 | -0.02 – 0.33 | 0.092 | -0.25 | -1.41 – 0.93 | 0.683 |
| >1 vs. mid | 0.11 | -0.07 – 0.29 | 0.235 | -0.26 | -1.43 – 0.91 | 0.665 |
| **Precipitation** |  |  |  |  |  |  |
| All Season ^b^ |  |  |  |  |  |  |
| <-0.5 vs. mid | -0.04 | -0.17 – 0.10 | 0.614 | 0.01 | -0.68 – 0.68 | 0.987 |
| >0.5 vs. mid | -0.04 | -0.14 – 0.05 | 0.386 | -0.06 | -0.56 – 0.43 | 0.801 |
| Spring |  |  |  |  |  |  |
| <-0.30 vs mid | -0.10 | -0.27 – 0.06 | 0.223 | -0.13 | -0.68 – 0.42 | 0.646 |
| >0.5 vs mid | -0.24 | -0.50 – 0.03 | 0.084 | 0.03 | -0.87 – 0.93 | 0.945 |
| Summer |  |  |  |  |  |  |
| <-0.5 vs. mid | 0.05 | -0.02 – 0.13 | 0.175 | 0.40 | 0.00 – 0.79 | 0.047* |
| >0.5 vs. mid | -0.04 | -0.12 – 0.04 | 0.371 | 0.40 | -0.02 – 0.81 | 0.061 |
| Fall |  |  |  |  |  |  |
| <-0.5 vs. mid | 0.07 | -0.25 – 0.39 | 0.665 | -0.85 | -3.70 – 2.00 | 0.557 |
| >0.5 vs. mid | 0.07 | -0.09 – 0.23 | 0.369 | -0.22 | -1.65 – 1.21 | 0.763 |
| Winter |  |  |  |  |  |  |
| <-0.5 vs. mid | -0.02 | -0.29 – 0.25 | 0.894 | -0.34 | -2.14 – 1.46 | 0.712 |
| >0.5 vs. mid | -0.24 | -0.43 - -0.05 | 0.012* | -0.15 | -1.42 – 1.11 | 0.811 |
| **Solar Radiation** |  |  |  |  |  |  |
| All Season ^b^ |  |  |  |  |  |  |
| <-1.5 vs. mid | 0.02 | -0.11 – 0.14 | 0.798 | -0.77 | -1.41 - -0.14 | 0.017* |
| >0 vs. mid | -0.12 | -0.19 - -0.05 | 0.001* ^d^ | -0.74 | -1.09 - -0.39 | <0.001* ^d^ |
| Spring |  |  |  |  |  |  |
| <-0.5 vs mid | 0.39 | 0.16 – 0.62 | 0.001* | -0.49 | -1.20 – 0.23 | 0.183 |
| >0.5 vs mid | -0.37 | -0.62 - -0.11 | 0.004* | -0.26 | -1.09 – 0.58 | 0.544 |
| Summer |  |  |  |  |  |  |
| <-1 vs. mid | -0.07 | -0.16 – 0.02 | 0.129 | 0.03 | -0.47 – 0.54 | 0.903 |
| >1 vs. mid | -0.21 | -0.27 - -0.14 | <0.001* | -0.14 | -0.48 – 0.19 | 0.396 |
| Fall |  |  |  |  |  |  |
| <-1.5 vs. mid | 0.03 | -0.16 – 0.21 | 0.793 | -1.24 | -2.92 – 0.43 | 0.146 |
| >0 vs. mid | -0.01 | -0.10 – 0.08 | 0.795 | -1.49 | -2.39 - -0.61 | 0.001* |
| Winter |  |  |  |  |  |  |
| <-1.5 vs. mid | -0.11 | -0.31 – 0.10 | 0.317 | -0.78 | -2.18 – 0.62 | 0.276 |
| >0 vs. mid | 0.26 | 0.13 – 0.39 | <0.001* ^d^ | -1.07 | -1.94 - -0.20 | 0.016* ^d^ |

β = Beta coefficient

CI = Confident Interval

***** p-value < 0.05

^a^ The beta coefficient, 95% confidence interval and p-value presented for each control variable correspond with the linear regression models assessing the impact of each on the mean percent of time indoors away from home (left) and 12-day lagged COVID-19 hospital admissions (right), including an auto-correlation term indicating yesterday’s response variable value

^b^ Models that were not stratified by season instead included season as a covariate to account for season as a confounder

^c^ The beta coefficient, 95% confidence interval and p-value presented for each independent weather variable correspond with the adjusted models assessing the impact of each treatment variable on the mean percent of time indoors away from home (left) and 12-day lagged COVID-19 hospital admissions (right), controlling for holidays and weekends, the stay-at-home order, increasing Colorado hospitalizations, as well as an auto-correlation term indicating yesterday’s response variable’s value

^d^ Weather variables that were associated with both mean percent of time spent indoors away-from-home and 12-day lagged hospitalizations are highlighted in gray, as these criteria was used to determine which variables to assess in the mediation analysis

**Supplemental Table S2: Sensitivity analysis results of the linear regression results using categorical weather conditions on time at home and COVID hospitalizations**

|  | **Daily county-mean percent of time spent at home (indoors or out)** | | | **12-day lagged hospitalization admissions (mean-centered by county-season)** | | |
| --- | --- | --- | --- | --- | --- | --- |
| **Control variables** ^a^ | **β** | **95% CI** | **p-value** | **β** | **95 % CI** | **p-value** |
| **Minimum temperature** |  |  |  |  |  |  |
| **All season** ^b^ |  |  |  |  |  |  |
| **Control variables** ^a^ | **β** | **95 % CI** | **p-value** | **β** | **95 % CI** | **p-value** |
| **Weekend/holiday (yes/no)** |  |  |  |  |  |  |
| All Seasons ^b^ | 1.01 | 0.89 – 1.14 | <0.001* | -0.22 | -0.56 – 0.12 | 0.201 |
| Spring | 1.46 | 1.20 – 1.73 | <0.001* | 0.06 | -0.51 – 0.63 | 0.835 |
| Summer | 0.34 | 1.15 – 0.53 | <0.001* | 0.28 | -0.03 – 0.60 | 0.080 |
| Fall | 0.60 | 0.36 – 0.85 | <0.001* | -0.24 | -1.09 – 0.62 | 0.584 |
| Winter | 1.44 | 1.17 – 1.71 | <0.001* | -0.67 | -1.49 – 0.15 | 0.109 |
| **Stay-at-home order (yes/no)** |  |  |  |  |  |  |
| All Seasons ^b^ | 1.00 | 0.69 – 1.31 | <0.001* | 0.19 | -0.32 – 0.69 | 0.469 |
| Spring | 0.46 | 0.08 – 0.85 | 0.019* | 0.28 | -0.26 – 0.82 | 0.305 |
| Summer | NA | NA | NA | NA | NA | NA |
| Fall | NA | NA | NA | NA | NA | NA |
| Winter | NA | NA | NA | NA | NA | NA |
| **Rising Colorado hospitalizations (yes/no)** |  |  |  |  |  |  |
| All Seasons ^b^ | 0.27 | 0.13 – 0.41 | <0.001* | 1.15 | 0.81 – 1.49 | <0.001* |
| Spring | 0.54 | 0.25 – 0.84 | <0.001* | 1.17 | 0.58 – 1.76 | <0.001* |
| Summer | -0.06 | -0.25 – 0.14 | 0.566 | 0.27 | -0.08 – 0.62 | 0.131 |
| Fall | 0.19 | -0.05 – 0.43 | 0.118 | 2.50 | 1.55 – 3.45 | <0.001* |
| Winter | 0.82 | 0.45 – 1.19 | <0.001* | 1.58 | 0.61 – 2.55 | 0.001* |
| **Categorical weather conditions** ^c^ | **β** | **95% CI** | **p-value** | **β** | **95 % CI** | **p-value** |
| **Minimum temperature** |  |  |  |  |  |  |
| **All season** ^b^ |  |  |  |  |  |  |
| <-0.5 vs. mid | 0.09 | -0.05 – 0.23 | 0.225 | 0.14 | -0.25 – 0.53 | 0.490 |
| >0.5 vs. mid | -0.08 | -0.22 – 0.05 | 0.239 | -0.41 | -0.78 - -0.03 | 0.034* |
| Spring |  |  |  |  |  |  |
| <-0.5 vs. mid | -0.04 | -0.32 – 0.24 | 0.782 | -0.18 | -0.78 – 0.42 | 0.557 |
| >0.5 vs. mid | 0.04 | -0.30 – 0.39 | 0.816 | -0.23 | -0.96 – 0.49 | 0.526 |
| Summer |  |  |  |  |  |  |
| <-0.5 vs. mid | 0.13 | -0.11 – 0.38 | 0.285 | -0.35 | -0.77 – 0.07 | 0.102 |
| >0.5 vs. mid | -0.22 | -0.41 - -0.03 | 0.022* | -0.14 | -0.47 – 0.19 | 0.394 |
| Fall |  |  |  |  |  |  |
| <-1 vs. mid | 0.86 | 0.51 – 1.20 | <0.001* | 0.61 | -0.56 – 1.78 | 0.306 |
| >1 vs. mid | -0.15 | -0.47 – 0.16 | 0.342 | -0.03 | -1.12 – 1.06 | 0.959 |
| Winter |  |  |  |  |  |  |
| <-1 vs. mid | 0.87 | 0.56 – 1,18 | <0.001* | 0.26 | -0.69 – 1.21 | 0.589 |
| >1 vs. mid | 0.15 | -0.21 – 0.51 | 0.417 | 1.19 | -0.01 – 2.40 | 0.053 |
| **Maximum temperature** |  |  |  |  |  |  |
| All season ^b^ |  |  |  |  |  |  |
| <-1 vs. mid | 0.52 | 0.36 – 0.67 | <0.001* | -0.19 | -0.62 – 0.23 | 0.374 |
| >1 vs. mid | -0.04 | -0.21 – 0.13 | 0.633 | -0.28 | -0.74 – 0.19 | 0.241 |
| Spring |  |  |  |  |  |  |
| <-1 vs mid | 0.54 | 0.23 – 0.86 | 0.001* ^d^ | -0.73 | -1.41 - -0.04 | 0.037* ^d^ |
| >1 vs mid | 0.19 | -0.27 – 0.64 | 0.421 | -0.62 | -1.58 – 0.33 | 0.201 |
| Summer |  |  |  |  |  |  |
| <-1 vs. mid | 0.23 | -0.03 – 0.48 | 0.080 | -0.48 | -0.94 - -0.02 | 0.039* |
| >1 vs. mid | -0.70 | -0.93 - -0.47 | <0.001* | -0.18 | -0.59 – 0.23 | 0.390 |
| Fall |  |  |  |  |  |  |
| <-1 vs. mid | 1.10 | 0.78 – 1.42 | <0.001* | 0.32 | -0.78 – 1.41 | 0.567 |
| >1 vs. mid | -0.14 | -0.46 – 0.18 | 0.389 | -0.28 | -1.42 – 0.86 | 0.632 |
| Winter |  |  |  |  |  |  |
| <-1 vs. mid | 0.88 | 0.58 – 1.18 | <0.001* | 0.40 | -0.59 – 1.39 | 0.431 |
| >1 vs. mid | -0.08 | -0.43 – 0.27 | 0.653 | 0.23 | -0.91 – 1.37 | 0.691 |
| **Minimum Relative Humidity** |  |  |  |  |  |  |
| All season ^b^ |  |  |  |  |  |  |
| <-1 vs. mid | 0.06 | -0.14 – 0.26 | 0.588 | -0.17 | -0.73 – 0.39 | 0.557 |
| >1 vs. mid | 0.75 | 0.59 – 0.91 | <0.001* | -0.27 | -0.72 – 0.18 | 0.238 |
| Spring |  |  |  |  |  |  |
| <-0.5 vs. mid | 0.13 | -0.13 – 0.38 | 0.328 | -0.03 | -0.60 – 0.55 | 0.926 |
| >1 vs. mid | 1.12 | 0.79 – 1.45 | <0.001* | -0.65 | -1.40 – 0.10 | 0.089 |
| Summer |  |  |  |  |  |  |
| <-0.5 vs. mid | -0.28 | -0.45 - -0.10 | 0.002** | 0.12 | -0.19 – 0.44 | 0.439 |
| >1 vs. mid | 0.20 | -0.05 – 0.45 | 0.118 | 0.02 | -0.43 – 0.47 | 0.930 |
| Fall |  |  |  |  |  |  |
| <-0.5 vs. mid | 0.02 | -0.22 – 0.26 | 0.866 | -0.05 | -0.90 – 0.79 | 0.899 |
| >1 vs. mid | 1.30 | 0.96 – 1.64 | <0.001* | 0.39 | -0.86 – 1.64 | 0.540 |
| Winter |  |  |  |  |  |  |
| <-1 vs. mid | -0.07 | -0.48 – 0.34 | 0.746 | -1.04 | -2.36 – 0.27 | 0.121 |
| >1 vs. mid | 0.79 | 0.44 – 1.13 | <0.001* | -0.01 | -1.11 – 1.09 | 0.985 |
| **Maximum relative humidity** |  |  |  |  |  |  |
| All Season ^b^ |  |  |  |  |  |  |
| <-1 vs. mid | 0.09 | -0.07 – 0.24 | 0.274 | -0.42 | -0.85 – 0.12 | 0.057 |
| >1 vs. mid | 0.44 | 0.30 – 0.59 | <0.001* | -0.36 | -0.76 – 0.04 | 0.081 |
| Spring |  |  |  |  |  |  |
| <-1 vs. mid | 0.42 | 0.10 – 0.74 | 0.009* | -0.14 | -0.84 – 0.57 | 0.706 |
| >1 vs. mid | 0.73 | 0.44 – 1.03 | <0.001* | -0.14 | -0.80 – 0.53 | 0.683 |
| Summer |  |  |  |  |  |  |
| <-0.5 vs. mid | -0.28 | -0.46 - -0.10 | 0.002* | 0.01 | -0.31 – 0.33 | 0.963 |
| >1 vs. mid | 0.01 | -0.23 – 0.24 | 0.958 | 0.00 | -0.42 – 0.42 | 0.999 |
| Fall |  |  |  |  |  |  |
| <-0.5 vs. mid | -0.06 | -0.32 – 0.19 | 0.638 | -0.08 | -0.96 – 0.80 | 0.856 |
| >1 vs. mid | 0.57 | 0.27 – 0.88 | <0.001* | -0.02 | -1.07 – 1.04 | 0.974 |
| Winter |  |  |  |  |  |  |
| <-1 vs. mid | 0.06 | -0.32 – 0.44 | 0.764 | -1.00 | -2.23 – 0.24 | 0.113 |
| >1 vs. mid | 0.73 | 0.42 – 1.04 | <0.001* | -0.45 | -1.44 – 0.54 | 0.376 |
| **Minimum absolute humidity** |  |  |  |  |  |  |
| All Season ^b^ |  |  |  |  |  |  |
| <-1 vs. mid | -0.23 | -0.40 – -0.06 | 0.010* | 0.08 | -0.40 – 0.56 | 0.740 |
| >1 vs. mid | 0.37 | 0.20 – 0.54 | <0.001* | -0.31 | -0.77 – 0.15 | 0.183 |
| Spring |  |  |  |  |  |  |
| <-1 vs mid | -0.28 | -0.63 – 0.07 | 0.117 | 0.45 | -0.33 – 1.22 | 0.259 |
| >1 vs mid | 1.01 | 0.64 – 1.38 | <0.001* | 0.07 | -0.76 – 0.90 | 0.865 |
| Summer |  |  |  |  |  |  |
| <-1 vs. mid | 0.05 | -0.20 – 0.30 | 0.681 | -0.03 | -0.48 – 0.41 | 0.892 |
| >1 vs. mid | 0.18 | -0.05 – 0.40 | 0.120 | -0.16 | -0.55 – 0.22 | 0.401 |
| Fall |  |  |  |  |  |  |
| <-1 vs. mid | -0.26 | -0.64 – 0.12 | 0.186 | 0.68 | -0.66 – 2.01 | 0.320 |
| >1 vs. mid | 0.18 | -0.17 – 0.54 | 0.309 | 0.07 | -1.14 – 1.29 | 0.910 |
| Winter |  |  |  |  |  |  |
| <-1 vs. mid | -0.35 | -0.72 – 0.03 | 0.069 | -1.28 | -2.50 - -0.07 | 0.039* |
| >1 vs. mid | 0.40 | -0.00 – 0.80 | 0.051 | -0.43 | -1.70 – 0.84 | 0.509 |
| **Maximum absolute humidity** |  |  |  |  |  |  |
| All Season ^b^ |  |  |  |  |  |  |
| <-1 vs. mid | 0.31 | 0.15 – 0.47 | <0.001* | 0.07 | -0.37 – 0.50 | 0.763 |
| >1 vs. mid | -0.08 | -0.25 – 0.09 | 0.364 | -0.11 | -0.58 – 0.36 | 0.657 |
| Spring |  |  |  |  |  |  |
| <-1 vs mid | 0.55 | 0.21 – 0.89 | 0.002* ^d^ | -1.00 | -1.74 – -0.26 | 0.008* ^d^ |
| >1 vs mid | 0.33 | -0.12 – 0.78 | 0.147 | -0.12 | -1.07 – 0.84 | 0.810 |
| Summer |  |  |  |  |  |  |
| <-1 vs. mid | -0.13 | -0.35 – 0.09 | 0.233 | 0.01 | -0.38 – 0.40 | 0.963 |
| >1 vs. mid | -0.37 | -0.58 – -0.15 | 0.001* | 0.00 | -0.38 – 0.38 | 0.993 |
| Fall |  |  |  |  |  |  |
| <-1 vs. mid | 1.41 | 1.03 – 1.78 | <0.001* | 1.27 | -0.04 – 2.57 | 0.058 |
| >1 vs. mid | -0.21 | -0.52 – 0.11 | 0.197 | 0.23 | -0.92 – 1.37 | 0.698 |
| Winter |  |  |  |  |  |  |
| <-1 vs. mid | 0.43 | 0.11 – 0.75 | 0.008* | 0.05 | -0.99 – 1.08 | 0.931 |
| >1 vs. mid | -0.50 | -1.00 – 0.00 | 0.050* | 0.05 | -1.56 – 1.67 | 0.950 |
| **Wind Speed** |  |  |  |  |  |  |
| All Season ^b^ |  |  |  |  |  |  |
| <-1 vs. mid | 0.15 | -0.03 – 0.32 | 0.102 | -0.09 | -0.57 – 0.40 | 0.730 |
| >1 vs. mid | 0.07 | -0.09 – 0.24 | 0.376 | 0.39 | -0.06 – 0.84 | 0.093 |
| Spring |  |  |  |  |  |  |
| <-1 vs mid | -0.01 | -0.37 – 0.35 | 0.949 | 0.13 | -0.64 – 0.90 | 0.745 |
| >1 vs mid | -0.06 | -0.40 – 0.28 | 0.746 | 0.03 | -0.70 – 0.76 | 0.930 |
| Summer |  |  |  |  |  |  |
| <-1 vs. mid | -0.21 | -0.47 – 0.06 | 0.133 | -0.09 | -0.57 – 0.40 | 0.725 |
| >1 vs. mid | 0.38 | 0.13 – 0.64 | 0.003* | -0.40 | -0.83 – 0.04 | 0.074 |
| Fall |  |  |  |  |  |  |
| <-1 vs. mid | 0.61 | 0.26 – 0.96 | 0.001* | -0.03 | -1.26 – 1.21 | 0.966 |
| >1 vs. mid | 0.64 | 0.30 – 0.97 | <0.001* | 1.11 | -0.08 – 2.29 | 0.067 |
| Winter |  |  |  |  |  |  |
| <-1 vs. mid | -0.01 | -0.38 – 0.36 | 0.968 | -0.25 | -1.41 – 0.93 | 0.683 |
| >1 vs. mid | -0.41 | -0.78 - -0.03 | 0.034* | -0.26 | -1.43 – 0.91 | 0.665 |
| **Precipitation** |  |  |  |  |  |  |
| All Season ^b^ |  |  |  |  |  |  |
| <-0.5 vs. mid | 0.09 | -0.16 – 0.33 | 0.487 | 0.01 | -0.68 – 0.68 | 0.987 |
| >0.5 vs. mid | 0.50 | 0.32 – 0.68 | <0.001* | -0.06 | -0.56 – 0.43 | 0.801 |
| Spring |  |  |  |  |  |  |
| <-0.30 vs mid | -0.10 | -0.35 – 0.15 | 0.436 | -0.13 | -0.68 – 0.42 | 0.646 |
| >0.5 vs mid | 0.10 | 0.57 – 1.40 | <0.001 | 0.03 | -0.87 – 0.93 | 0.945 |
| Summer |  |  |  |  |  |  |
| <-0.5 vs. mid | -0.01 | -0.24 – 0.21 | 0.908 | 0.40 | 0.00 – 0.79 | 0.047* |
| >0.5 vs. mid | 0.16 | -0.07 – 0.40 | 0.172 | 0.40 | -0.02 – 0.81 | 0.061 |
| Fall |  |  |  |  |  |  |
| <-0.5 vs. mid | 0.34 | -0.47 – 1.15 | 0.406 | -0.85 | -3.70 – 2.00 | 0.557 |
| >0.5 vs. mid | 0.80 | 0.39 – 1.21 | <0.001* | -0.22 | -1.65 – 1.21 | 0.763 |
| Winter |  |  |  |  |  |  |
| <-0.5 vs. mid | 0.14 | -0.42 – 0.70 | 0.619 | -0.34 | -2.14 – 1.46 | 0.712 |
| >0.5 vs. mid | 0.68 | 0.28 – 1.08 | 0.001* | -0.15 | -1.42 – 1.11 | 0.811 |
| **Solar Radiation** |  |  |  |  |  |  |
| All Season ^b^ |  |  |  |  |  |  |
| <-1.5 vs. mid | 0.38 | 0.15 – 0.61 | 0.001* ^d^ | -0.77 | -1.41 - -0.14 | 0.017* ^d^ |
| >0 vs. mid | -0.16 | -0.28 - -0.04 | 0.012* ^d^ | -0.74 | -1.09 - -0.39 | <0.001* ^d^ |
| Spring |  |  |  |  |  |  |
| <-0.5 vs mid | 0.25 | -0.12 – 0.62 | 0.183 | -0.49 | -1.20 – 0.23 | 0.183 |
| >0.5 vs mid | 0.35 | -0.04 – 0.74 | 0.077 | -0.26 | -1.09 – 0.58 | 0.544 |
| Summer |  |  |  |  |  |  |
| <-1 vs. mid | 0.17 | -0.11 – 0.45 | 0.240 | 0.03 | -0.47 – 0.54 | 0.903 |
| >1 vs. mid | -0.29 | -0.47 - -0.10 | 0.002* | -0.14 | -0.48 – 0.19 | 0.396 |
| Fall |  |  |  |  |  |  |
| <-1.5 vs. mid | 0.51 | 0.05 – 0.98 | 0.030* | -1.24 | -2.92 – 0.43 | 0.146 |
| >0 vs. mid | -0.81 | -1.07 - -0.55 | <0.001* ^d^ | -1.49 | -2.39 - -0.61 | 0.001* ^d^ |
| Winter |  |  |  |  |  |  |
| <-1.5 vs. mid | 0.37 | -0.07 – 0.81 | 0.097 | -0.78 | -2.18 – 0.62 | 0.276 |
| >0 vs. mid | -0.43 | -0.70 - -0.15 | 0.002* ^d^ | -1.07 | -1.94 - -0.20 | 0.016* ^d^ |

β = Beta coefficient

CI = Confident Interval

***** p-value < 0.05

^a^ The beta coefficient, 95% confidence interval and p-value presented for each control variable correspond with the linear regression models assessing the impact of each on the mean percent of time at home (left) and 12-day lagged COVID-19 hospital admissions (right), including an auto-correlation term indicating yesterday’s response variable value

^b^ Models that were not stratified by season instead included season as a covariate to account for season as a confounder

^c^ The beta coefficient, 95% confidence interval and p-value presented for each independent weather variable correspond with the adjusted models assessing the impact of each treatment variable on the mean percent of time at home (left) and 12-day lagged COVID-19 hospital admissions (right), controlling for holidays and weekends, the stay-at-home order, increasing Colorado hospitalizations, as well as an auto-correlation term indicating yesterday’s response variable’s value

^d^ Weather variables that were associated with both mean percent of time at home and 12-day lagged hospitalizations are highlighted in gray, as these criteria was used to determine which variables to assess in the mediation analysis

**Supplemental Table S3. Sensitivity analysis detailing mediation results of categorical weather conditions and time at home as the mediator**

|  |  |  | **Estimate of the mediating effects of time at home on 12-day lagged COVID-19 hospital admissions** | | |
| --- | --- | --- | --- | --- | --- |
|  | **Treatment level** ^a^ | **Effect** | **β** | **95% CI** | **P-Value** |
| **All Seasons** |  |  |  |  |  |
| High solar radiation | >0 SD vs. -1.5 – 0 SD | Natural Indirect Effect | -0.02 | -0.05 – 0.00 | 0.090 |
|  | >0 SD vs. -1.5 – 0 SD | Natural Direct Effect | -0.76 | -1.08 - -0.43 | <0.001* |
|  | >0 SD vs. -1.5 – 0 SD | Total Effect | -0.78 | -1.10 - -0.45 | <0.001* |
| Low solar radiation | <0 SD vs. -1.5 – 0 SD | Natural Indirect Effect | 0.02 | -0.03 – 0.06 | 0.481 |
|  | <0 SD vs. -1.5 – 0 SD | Natural Direct Effect | -0.80 | -1.33 - -0.28 | 0.003* |
|  | <0 SD vs. -1.5 – 0 SD | Total Effect | -0.79 | -1.31 - -0.26 | 0.003* |
| **Spring** |  |  |  |  |  |
| Low maximum temperature | <-1 SD vs. -1 – 1 SD | Natural Indirect Effect | -0.01 | -0.06 – 0.04 | 0.604 |
|  | <-1 SD vs. -1 – 1 SD | Natural Direct Effect | -0.73 | -1.33 - -0.13 | 0.016* |
|  | <-1 SD vs. -1 – 1 SD | Total Effect | -0.74 | -1.35 - -0.14 | 0.016* |
| Low maximum absolute humidity | <-1 SD vs. -1 – 1 SD | Natural Indirect Effect | 0.00 | -0.13 – 0.06 | 0.883 |
|  | <-1 SD vs. -1 – 1 SD | Natural Direct Effect | -1.00 | -1.66 – -0.34 | 0.003* |
|  | <-1 SD vs. -1 – 1 SD | Total Effect | -1.00 | -1.67 - -0.32 | 0.004* |
| **Fall** |  |  |  |  |  |
| High solar radiation | >0 SD vs. -1.5 – 0 SD | Natural Indirect Effect | -0.23 | -0.47 – 0.02 | 0.069 |
|  | >0 SD vs. -1.5 – 0 SD | Natural Direct Effect | -0.97 | -1.88 - -0.06 | 0.036* |
|  | >0 SD vs. -1.5 – 0 SD | Total Effect | -1.20 | -2.00 - -0.39 | 0.004* |
| **Winter** |  |  |  |  |  |
| High solar radiation | >0 SD vs. -1.5 – 0 SD | Natural Indirect Effect | -0.14 | -0.30 – 0.02 | 0.084 |
|  | >0 SD vs. -1.5 – 0 SD | Natural Direct Effect | -0.76 | -1.65 – 0.12 | 0.092 |
|  | >0 SD vs. -1.5 – 0 SD | Total Effect | -0.90 | -1.75 - -0.05 | 0.038* |

β = Beta coefficient

CI = Confident Interval

***** p-value < 0.05

^a^ Seasonal weather conditions were categorized into three groups by examining Lowess plots between the weather variable and both the mediator (time at home) and outcome (12-day lagged hospital admissions) in this analysis. Linear regression analyses compared the association of “high” and “low” weather categories (versus the mid-range) on both the mediator and outcome. Those seasonal weather conditions were significantly associated with both are included in this table

**Supplemental Table S4. Sensitivity analysis detailing linear regression results for continuous weather conditions on time indoors and away-from-home and COVID hospitalizations**

|  | **Daily county-mean percent of time spent indoors, away-from-home** | | | **12-day lagged hospitalization admissions (mean-centered by county-season)** | | |
| --- | --- | --- | --- | --- | --- | --- |
| **Categorical weather conditions** ^a^ | **β** | **95 % CI** | **p-value** | **β** | **95 % CI** | **p-value** |
| **Minimum temperature** |  |  |  |  |  |  |
| **All Seasons** ^b^ | **-0.07** | **-0.11 - -0.04** | **<0.001*** ^c^ | **-0.18** | **-0.35 – -0.02** | **0.029*** ^c^ |
| Spring | -0.21 | -0.30 – -0.12 | <0.001* | 0.07 | -0.23 – 0.37 | 0.643 |
| Summer | -0.00 | -0.03 – 0.02 | 0.733 | 0.02 | -0.13 – 0.17 | 0.773 |
| Fall | 0.01 | -0.04 – 0.06 | 0.659 | -0.36 | -0.78 – 0.07 | 0.102 |
| Winter | 0.11 | 0.05 – 0.18 | <0.001* | -0.04 | -0.47 – 0.38 | 0.836 |
| **Maximum temperature** |  |  |  |  |  |  |
| **All Seasons** ^b^ | **-0.05** | **-0.09 – -0.02** | **0.001*** | **-0.09** | **-0.25 – 0.07** | **0.264** |
| Spring | -0.15 | -0.24 – -0.06 | 0.001* | 0.20 | -0.09 – 0.48 | 0.177 |
| Summer | 0.01 | -0.02 – 0.04 | 0.558 | 0.05 | -0.09 – 0.19 | 0.505 |
| Fall | 0.02 | -0.03 – 0.07 | 0.390 | -0.34 | -0.76 – 0.08 | 0.111 |
| Winter | 0.12 | 0.06 – 0.18 | <0.001* | -0.19 | -0.60 – 0.22 | 0.359 |
| **Minimum relative humidity** |  |  |  |  |  |  |
| **All Seasons** ^b^ | **-0.00** | **-0.03 – 0.03** | **0.910** | **-0.07** | **-0.23 – 0.09** | **0.375** |
| Spring | 0.03 | -0.05 – 0.11 | 0.451 | -0.19 | -0.45 – 0.07 | 0.147 |
| Summer | -0.03 | -0.06 – 0.00 | 0.095 | -0.03 | -0.18 – 0.13 | 0.729 |
| Fall | -0.03 | -0.08 – 0.01 | 0.129 | 0.14 | -0.25 – 0.53 | 0.472 |
| Winter | -0.14 | -0.20 - -0.08 | <0.001* | 0.13 | -0.28 – 0.55 | 0.527 |
| **Maximum relative humidity** |  |  |  |  |  |  |
| **All Seasons** ^b^ | **0.02** | **-0.01 – 0.05** | **0.150** | **-0.02** | **-0.18 – 0.13** | **0.765** |
| Spring | 0.12 | 0.04 – 0.20 | 0.003* | -0.02 | -0.28 – 0.24 | 0.875 |
| Summer | -0.01 | -0.04 – 0.02 | 0.416 | 0.01 | -0.14 – 0.16 | 0.917 |
| Fall | -0.03 | -0.07 – 0.02 | 0.242 | 0.05 | -0.35 – 0.44 | 0.814 |
| Winter | -0.11 | -0.17 - -0.05 | <0.001* | 0.12 | -0.28 – 0.52 | 0.551 |
| **Minimum absolute humidity** |  |  |  |  |  |  |
| **All Seasons** ^b^ | **-0.05** | **-0.08 - -0.01** | **0.006*** | **-0.14** | **-0.30 – 0.01** | **0.075** |
| Spring | -0.08 | -0.17 – 0.01 | 0.088 | -0.16 | -0.45 – 0.13 | 0.269 |
| Summer | -0.04 | -0.07 – -0.01 | 0.003* | -0.05 | -0.19 – 0.09 | 0.498 |
| Fall | -0.01 | -0.05 – 0.04 | 0.765 | 0.05 | -0.36 – 0.45 | 0.826 |
| Winter | -0.09 | -0.16 - -0.03 | 0.006* | 0.11 | -0.33 – 0.56 | 0.620 |
| **Maximum absolute humidity** |  |  |  |  |  |  |
| **All Seasons** ^b^ | **-0.06** | **-0.09 - -0.02** | **0.001*** | **-0.15** | **-0.31 – 0.02** | **0.084** |
| Spring | -0.09 | -0.20 – 0.02 | 0.106 | 0.21 | -0.11 – 0.53 | 0.201 |
| Summer | -0.02 | -0.04 – 0.01 | 0.161 | 0.03 | -0.10 – 0.17 | 0.646 |
| Fall | -0.02 | -0.07 – 0.04 | 0.519 | -0.38 | -0.85 – 0.10 | 0.121 |
| Winter | 0.04 | -0.02 – 0.11 | 0.187 | -0.14 | -0.58 – 0.30 | 0.545 |
| **Mean shortwave radiation** |  |  |  |  |  |  |
| **All Seasons** ^b^ | **-0.09** | **-0.13 – -0.06** | **<0.001*** | **-0.17** | **-0.34 - -0.00** | **0.056** |
| Spring | -0.27 | -0.37 – -0.16 | <0.001* | 0.08 | -0.22 – 0.39 | 0.581 |
| Summer | -0.08 | -0.11 – 0.05 | <0.001* | -0.05 | -0.22 – 0.12 | 0.547 |
| Fall | -0.01 | -0.05 – 0.03 | 0.598 | -0.33 | -0.73 – 0.07 | 0.106 |
| Winter | 0.14 | 0.08 – 0.21 | <0.001* | -0.43 | -0.89 – 0.03 | 0.065 |
| **Total precipitation** |  |  |  |  |  |  |
| **All Seasons** ^b^ | **-0.01** | **-0.04 – 0.02** | **0.548** | **0.03** | **-0.13 – 0.19** | **0.710** |
| Spring | -0.11 | -0.20 – 0.02 | 0.013* | 0.13 | -0.17 – 0.43 | 0.393 |
| Summer | -0.02 | -0.04 – 0.01 | 0.120 | 0.10 | -0.03 – 0.23 | 0.122 |
| Fall | 0.00 | -0.04 – 0.05 | 0.917 | 0.05 | -0.35 – 0.46 | 0.788 |
| Winter | -0.09 | -0.15 – -0.02 | 0.007* | -0.07 | -0.49 – 0.36 | 0.758 |
| **Mean wind velocity** |  |  |  |  |  |  |
| **All Seasons** ^b^ | **0.01** | **-0.02 – 0.04** | **0.543** | **0.09** | **-0.06 – 0.25** | **0.253** |
| Spring | 0.05 | -0.03 – 0.13 | 0.257 | -0.10 | -0.36 – 0.17 | 0.480 |
| Summer | -0.04 | -0.06 – -0.01 | 0.010* | -0.03 | -0.18 – 0.11 | 0.653 |
| Fall | -0.04 | -0.08 – 0.01 | 0.138 | 0.05 | -0.38 – 0.48 | 0.817 |
| Winter | -0.01 | -0.07 – 0.06 | 0.820 | -0.03 | -0.45 – 0.39 | 0.879 |

β = Beta coefficient

CI = Confident Interval

***** p-value < 0.05

^a^ The beta coefficient, 95% confidence interval and p-value presented for each continuous independent weather variable correspond with the adjusted models assessing the impact of each treatment variable on the mean percent of time indoors away from home (left) and 12-day lagged COVID-19 hospital admissions (right), controlling for holidays and weekends, the stay-at-home order, increasing Colorado hospitalizations, as well as an auto-correlation term indicating yesterday’s response variable’s value

^b^ Models that were not stratified by season instead included season as a covariate to account for season as a confounder

^c^ Weather variables that were associated with both mean percent of time spent indoors away-from-home and 12-day lagged hospitalizations are highlighted in gray, as these criteria was used to determine which variables to assess in the mediation analysis

**Supplemental Table S5. Sensitivity analysis results for the mediation models using continuous weather variables and time indoors away-from-home as the mediator**

|  |  |  | **Estimating the mediating effects of time spent indoors away-from-home on 12-day lagged COVID hospital admissions** | | |
| --- | --- | --- | --- | --- | --- |
|  | **Treatment level** ^a^ | **Effect** | **β** | **95% CI** | **P-Value** |
| **All Seasons** |  |  |  |  |  |
| Low minimum temperature | -1 SD vs. mean | Natural Indirect Effect | -0.01 | -0.01 – 0.00 | 0.103 |
|  | -1 SD vs. mean | Natural Direct Effect | 0.20 | 0.05 – 0.37 | 0.013* |
|  | -1 SD vs. mean | Total Effect | 0.20 | 0.04 – 0.36 | 0.015* |
| High minimum temperature | +1 SD vs. mean | Natural Indirect Effect | 0.01 | -0.00 – 0.02 | 0.137 |
|  | +1 SD vs. mean | Natural Direct Effect | -0.20 | -0.37 – -0.04 | 0.013* |
|  | +1 SD vs. mean | Total Effect | -0.20 | -0.36 – -0.03 | 0.017* |

β = Beta coefficient

CI = Confident Interval

***** p-value < 0.05

^a^ Seasonal weather conditions were included in models as continuous standardized measures within the mediation models. The treatment group was defined as ±1 SD, while the control group was defined as the mean.

**Supplemental Table S6. Sensitivity analysis detailing linear regression results of continuous weather conditions on time at home, and on hospitalizations**

|  | **Percent of time at home**  **(mean-centered by county season)** | | | **12-day lagged hospitalization admissions (mean-centered by county-season)** | | |
| --- | --- | --- | --- | --- | --- | --- |
| **Continuous weather conditions** ^a^ | **β** | **95 % CI** | **p-value** | **β** | **95 % CI** | **p-value** |
| **Minimum temperature** |  |  |  |  |  |  |
| **All Seasons** ^b^ | **-0.09** | **-0.15 - -0.03** | **0.004*** ^c^ | **-0.18** | **-0.35 – -0.02** | **0.029*** ^c^ |
| Spring | 0.03 | -0.11 – 0.17 | 0.674 | 0.07 | -0.23 – 0.37 | 0.643 |
| Summer | -0.22 | -0.31 – -0.12 | <0.001* | 0.02 | -0.13 – 0.17 | 0.773 |
| Fall | -0.43 | -0.56 – -0.29 | <0.001* | -0.36 | -0.78 – 0.07 | 0.102 |
| Winter | -0.30 | -0.43 - -0.16 | <0.001* | -0.04 | -0.47 – 0.38 | 0.836 |
| **Maximum temperature** |  |  |  |  |  |  |
| **All Seasons** ^b^ | **-0.17** | **-0.23 – -0.12** | **<0.001*** | **-0.09** | **-0.25 – 0.07** | **0.264** |
| Spring | -0.15 | -0.29 – -0.02 | 0.027* | 0.20 | -0.09 – 0.48 | 0.177 |
| Summer | -0.23 | -0.31 - -0.14 | <0.001* | 0.05 | -0.09 – 0.19 | 0.505 |
| Fall | -0.50 | -0.63 – -0.38 | <0.001* | -0.34 | -0.76 – 0.08 | 0.111 |
| Winter | -0.31 | -0.44 – -0.18 | <0.001* | -0.19 | -0.60 – 0.22 | 0.359 |
| **Minimum relative humidity** |  |  |  |  |  |  |
| **All Seasons** ^b^ | **0.27** | **0.21 – 0.32** | **<0.001*** | **-0.07** | **-0.23 – 0.09** | **0.375** |
| Spring | 0.37 | 0.25 – 0.48 | <0.001* | -0.19 | -0.45 – 0.07 | 0.147 |
| Summer | 0.18 | 0.09 – 0.27 | <0.001* | -0.03 | -0.18 – 0.13 | 0.729 |
| Fall | 0.39 | 0.28 – 0.51 | <0.001* | 0.14 | -0.25 – 0.53 | 0.472 |
| Winter | 0.33 | 0.20 – 0.46 | <0.001* | 0.13 | -0.28 – 0.55 | 0.527 |
| **Maximum relative humidity** |  |  |  |  |  |  |
| **All Seasons** ^b^ | **0.15** | **0.09 – 0.20** | **<0.001*** | **-0.02** | **-0.18 – 0.13** | **0.765** |
| Spring | 0.17 | 0.05 – 0.29 | 0.005* | -0.02 | -0.28 – 0.24 | 0.875 |
| Summer | 0.13 | 0.04 – 0.21 | 0.003* | 0.01 | -0.14 – 0.16 | 0.917 |
| Fall | 0.24 | 0.12 – 0.36 | <0.001* | 0.05 | -0.35 – 0.44 | 0.814 |
| Winter | 0.22 | 0.10 – 0.35 | 0.001* | 0.12 | -0.28 – 0.52 | 0.551 |
| **Minimum absolute humidity** |  |  |  |  |  |  |
| **All Seasons** ^b^ | **0.17** | **0.11 – 0.22** | **<0.001*** | **-0.14** | **-0.30 – 0.01** | **0.075** |
| Spring | 0.40 | 0.27 – 0.53 | <0.001* | -0.16 | -0.45 – 0.13 | 0.269 |
| Summer | 0.07 | -0.01 – 0.16 | 0.095 | -0.05 | -0.19 – 0.09 | 0.498 |
| Fall | 0.11 | -0.01 – 0.23 | 0.062 | 0.05 | -0.36 – 0.45 | 0.826 |
| Winter | 0.17 | 0.03 – 0.31 | 0.017* | 0.11 | -0.33 – 0.56 | 0.620 |
| **Maximum absolute humidity** |  |  |  |  |  |  |
| **All Seasons** ^b^ | **-0.12** | **-0.18 - -0.06** | **<0.001*** | **-0.15** | **-0.31 – 0.02** | **0.084** |
| Spring | -0.07 | -0.23 – 0.09 | 0.390 | 0.21 | -0.11 – 0.53 | 0.201 |
| Summer | -0.13 | -0.21 – -0.05 | 0.001* | 0.03 | -0.10 – 0.17 | 0.646 |
| Fall | -0.46 | -0.60 – -0.32 | <0.001* | -0.38 | -0.85 – 0.10 | 0.121 |
| Winter | -0.24 | -0.38 – -0.11 | 0.001* | -0.14 | -0.58 – 0.30 | 0.545 |
| **Mean shortwave radiation** |  |  |  |  |  |  |
| **All Seasons** ^b^ | **-0.14** | **-0.20 – -0.08** | **<0.001*** | **-0.17** | **-0.34 - -0.00** | **0.056** |
| Spring | -0.09 | -0.25 – 0.07 | 0.264 | 0.08 | -0.22 – 0.39 | 0.581 |
| Summer | -0.11 | -0.21 – -0.02 | 0.017* | -0.05 | -0.22 – 0.12 | 0.547 |
| Fall | -0.51 | -0.63 – -0.39 | <0.001* | -0.33 | -0.73 – 0.07 | 0.106 |
| Winter | -0.31 | -0.45 – -0.16 | <0.001* | -0.43 | -0.89 – 0.03 | 0.065 |
| **Total precipitation** |  |  |  |  |  |  |
| **All Seasons** ^b^ | 0.19 | **0.13 – 0.24** | **<0.001*** | **0.03** | **-0.13 – 0.19** | **0.710** |
| Spring | 0.40 | 0.26 – 0.53 | <0.001* | 0.13 | -0.17 – 0.43 | 0.393 |
| Summer | 0.06 | -0.01 – 0.13 | 0.119 | 0.10 | -0.03 – 0.23 | 0.122 |
| Fall | 0.27 | 0.16 – 0.39 | <0.001* | 0.05 | -0.35 – 0.46 | 0.788 |
| Winter | 0.22 | 0.08 – 0.35 | 0.001* | -0.07 | -0.49 – 0.36 | 0.758 |
| **Mean wind velocity** |  |  |  |  |  |  |
| **All Seasons** ^b^ | **0.00** | **-0.05 – 0.06** | **0.903** | **0.09** | **-0.06 – 0.25** | **0.253** |
| Spring | -0.04 | -0.17 – 0.08 | 0.502 | -0.10 | -0.36 – 0.17 | 0.480 |
| Summer | 0.16 | 0.08 – 0.25 | <0.001* | -0.03 | -0.18 – 0.11 | 0.653 |
| Fall | 0.14 | 0.02 – 0.27 | 0.021* | 0.05 | -0.38 – 0.48 | 0.817 |
| Winter | -0.08 | -0.22 – 0.05 | 0.224 | -0.03 | -0.45 – 0.39 | 0.879 |

β = Beta coefficient

CI = Confident Interval

***** p-value < 0.05

^a^ The beta coefficient, 95% confidence interval and p-value presented for each continuous independent weather variable correspond with the adjusted models assessing the impact of each treatment variable on the mean percent of time indoors at home (left) and 12-day lagged COVID-19 hospital admissions (right), controlling for holidays and weekends, the stay-at-home order, increasing Colorado hospitalizations, as well as an auto-correlation term indicating yesterday’s response variable’s value

^b^ Models that were not stratified by season instead included season as a covariate to account for season as a confounder

^c^ Weather variables that were associated with both mean percent of time at home and 12-day lagged hospitalizations are highlighted in gray, as these criteria was used to determine which variables to assess in the mediation analysis

**Supplemental Table S7. Sensitivity analysis detailing mediation results assessing time at home as a mediator between continuous weather conditions and COVID hospital admissions**

|  |  |  | **Estimating the mediating effects of time spent at home on 12-day lagged COVID hospital admissions** | | |
| --- | --- | --- | --- | --- | --- |
|  | **Treatment level** ^a^ | **Effect** | **β** | **95% CI** | **P-Value** |
| **All Seasons** |  |  |  |  |  |
| Low minimum temperature | -1 SD vs. mean | Natural Indirect Effect | 0.01 | -0.00 – 0.01 | 0.113 |
|  | -1 SD vs. mean | Natural Direct Effect | 0.16 | 0.01 – 0.32 | 0.036* |
|  | -1 SD vs. mean | Total Effect | 0.17 | 0.02 – 0.32 | 0.031* |
| High minimum temperature | +1 SD vs. mean | Natural Indirect Effect | -0.01 | -0.02 – 0.00 | 0.103 |
|  | +1 SD vs. mean | Natural Direct Effect | -0.16 | -0.32 – -0.01 | 0.036* |
|  | +1 SD vs. mean | Total Effect | -0.17 | -0.33 – -0.02 | 0.027* |

β = Beta coefficient

CI = Confident Interval

***** p-value < 0.05

^a^ Seasonal weather conditions were included in models as continuous standardized measures within the mediation models. The treatment group was defined as ±1 SD, while the control group was defined as the mean.

**Supplemental Table S8. Sensitivity analysis detailing mediation results without hospitalization growth, using categorical weather conditions and time indoors away from home as the mediator**

|  |  |  | **With Hosp Growth** | | | **Without Hosp Growth** | | |
| --- | --- | --- | --- | --- | --- | --- | --- | --- |
|  | **Treatment level** ^a^ | **Effect** | **β** | **95% CI** | **p-value** | **β** | **95% CI** | **p-value** |
| **All-seasons** |  |  |  |  |  |  |  |  |
| High minimum temperature | >0.5 SD vs. -0.5 – 0.5 SD | Natural Indirect Effect | 0.02 | -0.01 – 0.04 | 0.144 | 0.01 | -0.01 – 0.03 | 0.324 |
|  | >0.5 SD vs. -0.5 – 0.5 SD | Natural Direct Effect | -0.46 | -0.82 – -0.11 | 0.011* | -0.54 | -0.90 - -0.18 | 0.003* |
|  | >0.5 SD vs. -0.5 – 0.5 SD | Total Effect | -0.45 | -0.80 – -0.10 | 0.013* | -0.53 | -0.88 - -0.18 | 0.003* |
| High solar radiation | >0 SD vs. -1.5 – 0 SD | Natural Indirect Effect | 0.03 | -0.00 – 0.07 | 0.054 | 0.02 | -0.01 – 0.04 | 0.184 |
|  | >0 SD vs. -1.5 – 0 SD | Natural Direct Effect | -0.88 | -1.23 - -0.53 | <0.001* | -1.00 | -1.35 - -0.64 | <0.001* |
|  | >0 SD vs. -1.5 – 0 SD | Total Effect | -0.85 | -1.19 - -0.51 | <0.001* | -0.98 | -1.33 - -0.63 | <0.001* |
| **Winter** |  |  |  |  |  |  |  |  |
| Low minimum absolute humidity | <-1 SD vs. -1 – 1 SD | Natural Indirect Effect | -0.10 | -0.35 – 0.16 | 0.462 | -0.11 | -0.29 – 0.07 | 0.248 |
|  | <-1 SD vs. -1 – 1 SD | Natural Direct Effect | -1.19 | -2.44 – 0.06 | 0.063 | -0.60 | -1.67 – 0.47 | 0.274 |
|  | <-1 SD vs. -1 – 1 SD | Total Effect | -1.29 | -2.59 – 0.01 | 0.052 | -0.71 | -1.84 – 0.43 | 0.223 |
| High solar radiation | >0 SD vs. -1.5 – 0 SD | Natural Indirect Effect | -0.09 | -0.26 – 0.08 | 0.300 | -0.16 | -0.35 – 0.03 | 0.092 |
|  | >0 SD vs. -1.5 – 0 SD | Natural Direct Effect | -0.95 | -1.82 - -0.09 | 0.030* | -0.91 | -1.69 – 0.01 | 0.054 |
|  | >0 SD vs. -1.5 – 0 SD | Total Effect | -1.04 | -1.87 - -0.21 | 0.014* | -0.87 | -1.83 - -0.17 | 0.019* |
| **Spring** |  |  |  |  |  |  |  |  |
| Low maximum temperature | <-1 SD vs. -1 – 1 SD | Natural Indirect Effect | 0.01 | -0.03 – 0.05 | 0.542 | -0.00 | -0.04 – 0.03 | 0.832 |
|  | <-1 SD vs. -1 – 1 SD | Natural Direct Effect | -0.78 | -1.42 - -0.13 | 0.018* | -0.77 | -1.43 - -0.11 | 0.022* |
|  | <-1 SD vs. -1 – 1 SD | Total Effect | -0.76 | -1.39 - -0.14 | 0.017* | -0.77 | -1.41 - -0.13 | 0.019* |

β = Beta coefficient

CI = Confident Interval

***** p-value < 0.05

^a^ Seasonal weather conditions were categorized into three groups by examining Lowess plots between the weather variable and both the mediator (time at home) and outcome (12-day lagged hospital admissions) in this analysis. Linear regression analyses compared the association of “high” and “low” weather categories (versus the mid-range) on both the mediator and outcome. Those seasonal weather conditions were significantly associated with both are included in this table

**Supplemental Table S9. Sensitivity analysis detailing mediation results without hospitalization growth, using categorical weather conditions and time at home as the mediator**

|  |  |  | **With Hosp Growth** | | | **Without Hosp Growth** | | |
| --- | --- | --- | --- | --- | --- | --- | --- | --- |
|  | **Treatment level** ^a^ | **Effect** | **β** | **95% CI** | **P-Value** | **β** | **95% CI** | **p-value** |
| **All Seasons** |  |  |  |  |  |  |  |  |
| High solar radiation | >0 SD vs. -1.5 – 0 SD | Natural Indirect Effect | -0.02 | -0.05 – 0.00 | 0.090 | -0.03 | -0.06 – 0.00 | 0.066 |
|  | >0 SD vs. -1.5 – 0 SD | Natural Direct Effect | -0.76 | -1.08 - -0.43 | <0.001* | -0.90 | -1.24 - -0.57 | <0.001* |
|  | >0 SD vs. -1.5 – 0 SD | Total Effect | -0.78 | -1.10 - -0.45 | <0.001* | -0.93 | -1.27 - -0.59 | <0.001* |
| Low solar radiation | <0 SD vs. -1.5 – 0 SD | Natural Indirect Effect | 0.02 | -0.03 – 0.06 | 0.481 | 0.02 | -0.02 – 0.06 | 0.362 |
|  | <0 SD vs. -1.5 – 0 SD | Natural Direct Effect | -0.80 | -1.33 - -0.28 | 0.003* | -0.86 | -1.40 - -0.32 | 0.002* |
|  | <0 SD vs. -1.5 – 0 SD | Total Effect | -0.79 | -1.31 - -0.26 | 0.003* | -0.93 | -1.27 - -0.59 | <0.001* |
| **Winter** |  |  |  |  |  |  |  |  |
| High solar radiation | >0 SD vs. -1.5 – 0 SD | Natural Indirect Effect | -0.14 | -0.30 – 0.02 | 0.084 | -0.17 | -0.33 - -0.01 | 0.041* |
|  | >0 SD vs. -1.5 – 0 SD | Natural Direct Effect | -0.76 | -1.65 – 0.12 | 0.092 | -0.65 | -1.51 – 0.22 | 0.143 |
|  | >0 SD vs. -1.5 – 0 SD | Total Effect | -0.90 | -1.75 - -0.05 | 0.038* | -0.81 | -1.66 – 0.03 | 0.060 |
| **Spring** |  |  |  |  |  |  |  |  |
| Low maximum temperature | <-1 SD vs. -1 – 1 SD | Natural Indirect Effect | -0.01 | -0.06 – 0.04 | 0.604 | 0.00 | -0.04 – 0.05 | 0.865 |
|  | <-1 SD vs. -1 – 1 SD | Natural Direct Effect | -0.73 | -1.33 - -0.13 | 0.016* | -0.79 | -1.39 - -0.18 | 0.011* |
|  | <-1 SD vs. -1 – 1 SD | Total Effect | -0.74 | -1.35 - -0.14 | 0.016* | -0.78 | -1.40 - -0.16 | 0.013* |
| Low maximum absolute humidity | <-1 SD vs. -1 – 1 SD | Natural Indirect Effect | 0.00 | -0.13 – 0.06 | 0.883 | 0.02 | -0.03 – 0.07 | 0.466 |
|  | <-1 SD vs. -1 – 1 SD | Natural Direct Effect | -1.00 | -1.66 – -0.34 | 0.003* | -1.06 | -1.73 - -0.40 | 0.002* |
|  | <-1 SD vs. -1 – 1 SD | Total Effect | -1.00 | -1.67 - -0.32 | 0.004* | -1.04 | -1.72 - -0.36 | 0.003* |
| **Fall** |  |  |  |  |  |  |  |  |
| High solar radiation | >0 SD vs. -1.5 – 0 SD | Natural Indirect Effect | -0.23 | -0.47 – 0.02 | 0.069 | -0.26 | -0.51 - -0.00 | 0.047* |
|  | >0 SD vs. -1.5 – 0 SD | Natural Direct Effect | -0.97 | -1.88 - -0.06 | 0.036* | -0.84 | -1.81 – 0.13 | 0.089 |
|  | >0 SD vs. -1.5 – 0 SD | Total Effect | -1.20 | -2.00 - -0.39 | 0.004* | -1.10 | -1.95 - -0.25 | 0.012* |

β = Beta coefficient

CI = Confident Interval

***** p-value < 0.05

^a^ Seasonal weather conditions were categorized into three groups by examining Lowess plots between the weather variable and both the mediator (time at home) and outcome (12-day lagged hospital admissions) in this analysis. Linear regression analyses compared the association of “high” and “low” weather categories (versus the mid-range) on both the mediator and outcome. Those seasonal weather conditions were significantly associated with both are included in this table
